# Causal links between inflammatory bowel disease and both psoriasis and psoriatic arthritis: a bidirectional two-sample Mendelian randomization study

**DOI:** 10.1101/2022.05.23.22275448

**Authors:** Dennis Freuer, Jakob Linseisen, Christa Meisinger

## Abstract

**Introduction:** Psoriasis (PsO), psoriatic arthritis (PsA) and inflammatory bowel disease (IBD), i.e. Crohn’s disease (CD) and ulcerative colitis (UC) are chronic systemic immune-mediated disorders affecting an increasing proportion of adults and children worldwide. Observational studies suggested an association between IBD and PsO and vice versa. However, so far it remains unclear whether a causal relationship exists.

**Methods:** To investigate the causal paths, a bidirectional two-sample Mendelian randomization (MR) study was conducted using summary statistics from genome-wide association studies (GWASs) including up to 463,372 Europeans. Total and direct effects were derived performing an iterative radial and robust inverse-variance weighted method within the univariable and multivariable MR setting, respectively. Causal estimates were verified using a validation IBD-sample, a series of pleiotropy-robust MR-methods, and sensitivity analyses based on PhenoScanner search in conjunction with network analysis.

**Results:** Genetically predicted IBD was associated with higher risk of PsO (pooled OR=1.10; 95% CI: (1.05; 1.15); P=1⋅10^−4^) and PsA (pooled OR=1.10; 95% CI: (1.04; 1.18); P=3⋅10^−3^). In contrast to UC, the CD subentity was related to PsO (OR=1.16; 95% CI: (1.12; 1.20); P=1⋅10^−14^) and PsA (OR=1.13; 95% CI: (1.06; 1.20); P=1⋅10^−4^). Regarding the reverse directions, no notable associations could be found.

**Conclusions:** This study supports a causal effect between IBD and PsO as well as PsA, but not vice versa. It seems that mainly CD and not UC is responsible for the causal impact of IBD on both psoriasis outcomes. These findings have implications for the management of IBD and psoriasis in clinical practice.

## Introduction

Psoriasis (PsO) is a chronic relapsing immune-mediated disease of the skin affecting approximately 2-3% of people worldwide (1). The prevalence ranges from 0.5% to 11.4% in adults and approximately 1.4% in children (2). PsO is caused by a constellation of environmental, immunogenic, and genetic factors (3), is chronic, incurable, and physically and emotionally debilitating (4–6). It is not only a disease of the skin, but a systemic disease that also affects other parts of the body, such as the joints, the cardiovascular system, and the central nervous system (7). About 30% of PsO patients suffer from psoriatic arthritis (PsA), a systemic inflammatory arthritis sharing common pathogenetic and immunologic features with PsO (8–11). Furthermore, now it is recognized that PsO is associated with other immune-mediated inflammatory conditions; in particular inflammatory bowel disease (IBD), including Crohn’s disease and ulcerative colitis as the main subtypes, appears to frequently co-occur with the disease (12–14). Evidence is accumulating that PsO and IBD share several genetic susceptibility loci and that the pathogenetic mechanisms underlying both diseases partially overlap (15–17).

Some previous cross-sectional and prospective epidemiological studies as well as meta-analysis reported an increased risk of IBD in patients with PsO and vice versa (18–22). However, so far, it remains unclear whether there is a causal relationship between these two inflammatory diseases because in observational studies bias due to confounding and reverse causation cannot be excluded. Therefore, in the present study, we performed a two-sample summary data MR analysis to investigate the causal effects of IBD including its subentities Crohn’s disease (CD) and ulcerative colitis (UC) on the risk of both PsO and PsA and vice versa.

## Methods

### Study design

To investigate the causal paths between IBD and PsO as well as PsA, we performed iteratively bidirectional two-sample Mendelian randomization (MR) analyses based on summary statistics of genome-wide association studies (GWASs). In an instrumental variable setting MR utilizes genetic variants as proxies for a specific modifiable risk factor to estimate and test for a causal effect with an outcome. The random allocation of genetic variants at the conception based on Mendel’s laws ensures the independency from any confounding factors and in this way mimics a randomized controlled trial. Moreover, if the core assumptions (relevance, independence, and exclusion restriction assumptions) are met, this study design overcomes reverse causation that may be an issue in observational studies. Details about the MR-design can be found elsewhere (23, 24).

### Study samples and measures

With regard to the evidence we used two different GWAS-datasets for IBD. The discovery sample included 12,882 clinically diagnosed IBD cases (21,770 controls) based on up to 15 studies considering the European population (25). Additionally, 5,956 diagnosed CD cases (14,927 controls) and 6,968 diagnosed UC cases (20,464 controls) were available from this cohort [Table 1]. Diagnosis was made by radiological, endoscopic and histopathological examinations. The validation sample consisted of 7,045 self-reported IBD cases (456,327 controls) from the UK Biobank cohort (26). A total of 5,621 PsO and 2,063 PsA cases were obtained from the FINNGEN Consortium and compared with 252,323 controls of European ancestry (27).

**Table 1.**
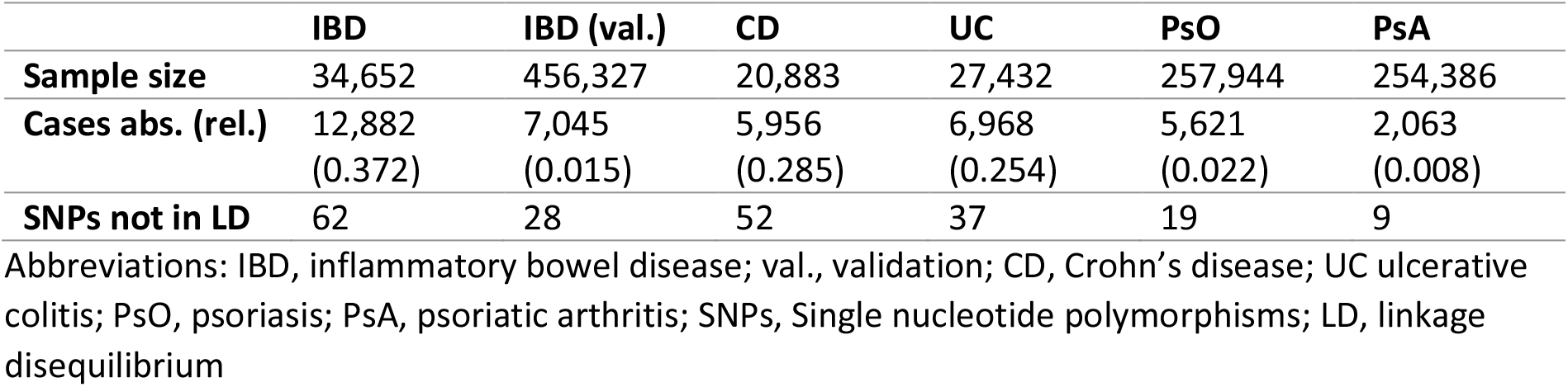
Description of samples

|  | **IBD** | **IBD (val.)** | **CD** | **UC** | **PsO** | **PsA** |
| --- | --- | --- | --- | --- | --- | --- |
| **Sample size** | 34,652 | 456,327 | 20,883 | 27,432 | 257,944 | 254,386 |
| **Cases abs. (rel.)** | 12,882<br>(0.372) | 7,045<br>(0.015) | 5,956<br>(0.285) | 6,968<br>(0.254) | 5,621<br>(0.022) | 2,063<br>(0.008) |
| **SNPs not in LD** | 62 | 28 | 52 | 37 | 19 | 9 |

Abbreviations: IBD, inflammatory bowel disease; val., validation; CD, Crohn's disease; UC ulcerative colitis; PsO, psoriasis; PsA, psoriatic arthritis; SNPs, Single nucleotide polymorphisms; LD, linkage disequilibrium

### Instruments selection

For each analysis we selected single nucleotide polymorphisms (SNPs) that were associated with the respective exposure at the genome-wide significance threshold *P* = 5 ⋅ 10^−8^. Where possible, we restricted the SNPs to an imputation score ≥ 0.8. To ensure independency, the genetic instruments underwent a PLINK clumping process. Regarding the European 1000 genomes reference panel, SNPs in linkage disequilibrium were pruned within a 10,000 kb window considering a minor allele frequency > 0.01 and a clumping cut-off *r*^2^ of 0.001. Within the harmonization step, we removed palindromic SNPs with a minor allele frequency > 0.42 and searched, if necessary, for proxy-SNPs in the outcome dataset using an *r*^2^ > 0.8.

Thus, starting with the number of independent SNPs reported in Table 1, 26 to 61 potential instruments for IBD and its subentities and 4 to 15 instrumental SNPs for the psoriasis phenotypes remained after the harmonization process [Table 2, Supplementary Table 1].

**Table 2.** Number of potential instruments used as starting basis for the analyses after the harmonization process

|  | **Psoriasis** |  | **Arthropathic psoriasis** |  |
| --- | --- | --- | --- | --- |
|  | Forward | Backward | Forward | Backward |
| **IBD** | 61 | 11 | 61 | 5 |
| **IBD (val)** | 26 | 15 | 26 | 6 |
| **CD** | 50 | 11 | 50 | 4 |
| **UC** | 34 | 12 | 34 | 5 |

Abbreviations: IBD, inflammatory bowel disease; val, validation; CD, Crohn's disease; UC ulcerative colitis; PsO, psoriasis; PsA, psoriatic arthritis

### Statistical analyses

The statistical analysis consists of 3 parts. First, univariable MR analyses were performed to investigate total effects. Second, multivariable MR analyses were conducted to distinguish the effects of either the IBD subentities or PsO and PsA and doing so, to calculate direct effects. Third, a series of sensitivity analyses were applied to assess the consistency of estimates and thus the plausibility of the MR core-assumptions. This was done by applying pleiotropy-robust MR methods as well as by excluding instruments that are likely to be associated with confounding factors of the respective exposure-outcome association.

#### Univariable MR

In detail, within the radial regression framework we used iteratively the inverse variance-weighted (IVW) method with modified second order weights as the main analysis. In each iteration step, outlier-SNPs were detected and removed based on the SNP-specific Cochran’s *Q* and Rucker’s *Q*′ statistics based on a type I error *α*_*Q*_ *= α*_*Q*′_ *=* 0.01 [Supplementary Table 2]. In addition, under a fixed effect model a pooled causal meta-analysis estimate was derived for both IBD samples.

#### Multivariable MR

Regarding the overlap of genetic instruments between both IBD subentities CD and UC as well as PsO and PsA, we performed bidirectional multivariable MR analyses to calculate the direct effects, that is, the influence on an outcome that can be directly attributed to the exposure of interest and not to another. As the main approach, the robust IVW method with multiplicative random effect was applied and heterogeneity was assessed using *Q* statistics.

#### Sensitivity analyses

To evaluate possible horizontal pleiotropy in case of invalid instruments, a broad range of methods, which are robust to specific patterns of heterogeneity, were applied to the final models. In the univariable case, these include the MR-Egger, weighted median, and weighted mode methods (28). Additionally, a many weak instrument analysis was done using the Robust Adjusted Profile Score (RAPS) with a robust loss function and consideration of overdispersion (29). The MR-PRESSO framework (30) was used to test for pleiotropy (global test), if necessary to correct the estimate by removing outliers (outlier test), and test for the distortion between both estimates. Directional pleiotropy in the final models was assessed using the radial MR-Egger intercept test and influential SNPs were investigated within a leave-one-out analysis as well as graphical evaluation of the radial and scatter (SNP-exposure vs. SNP-outcome associations) plots. In the multivariable setting MR-Egger, median, modified IVW, and MR-Lasso (31) methods were used as sensitivity analyses, whereas directional pleiotropy was tested with the MR-Egger intercept test.

As a further sensitivity analysis, we accounted for possible confounding factors of the exposure-outcome associations using following steps in both univariable and multivariable settings. We applied a PhenoScanner search (32, 33) to assess all known phenotypes related to the considered genetic instruments in our analyses. We then performed a network analysis to cluster all identified phenotypes into logical groups rather than just looking at individual phenotypes (e.g. obesity rather than body mass index or waist to hip ratio). Finally, we removed SNPs associated with one cluster per analysis and compared them with the causal estimates from the main approaches.

Based on a type I error *α =* 0.05, in this study evidence for a causal relationship was considered sufficient, if and only if, all methods used confirmed it with respect to heterogeneity statistics as well as consistent point estimates from sensitivity analyses. Estimates were presented as odds ratios (ORs) with 95% confidence intervals (CIs) and can be interpreted as the average change in the outcome per 2.72-fold increase in the prevalence of the respective binary exposure.

Analyses were done using the statistical software R (version 4.1.2). For the most of the part, the packages TwoSampleMR (version 0.5.6), RadialMR (version 1.0), MRPRESSO (version 1.0), MendelianRandomization (version 0.6.0), mr.raps (version 0.2), MVMR (version 0.3), data.table (version 1.14.2), dplyr (version 1.0.8), and ggplot2 (version 3.3.5) were used. Gephi (version 0.9) was used for the network analysis.

## Results

### Main results

#### Univariable MR

In the univariable case, main results were derived from the radial IVW method with modified second order weights after the last iteration and presented as ORs with 95% CIs. Genetically predicted IBD was positively related to both PsO (*OR*=1.09, CI: 1.04; 1.15, *P*= 1 ⋅ 10^−3^) and PsA (*OR*=1.09, CI: 1.01; 1.17, *P*= 2.6 ⋅ 10^−2^), which was confirmed by the validation sample [Figure 1]. The pooled meta-analysis estimates were (*OR*=1.10, CI: 1.05; 1.15, *P*= *5* ⋅ 10^−*5*^) for PsO and (*OR*=1.11, CI: 1.04; 1.18, *P*= 3 ⋅ 10^−3^) for PsA [Figure 2]. Subgroup analyses revealed that these associations could be attributed to CD (*ORs* between 1.13 and 1.16) but not UC (*ORs* between 1.01 and 1.03) [Figure 1]. There was no indication of causal effects in the reverse directions [Figure 1].

**Figure 1.**
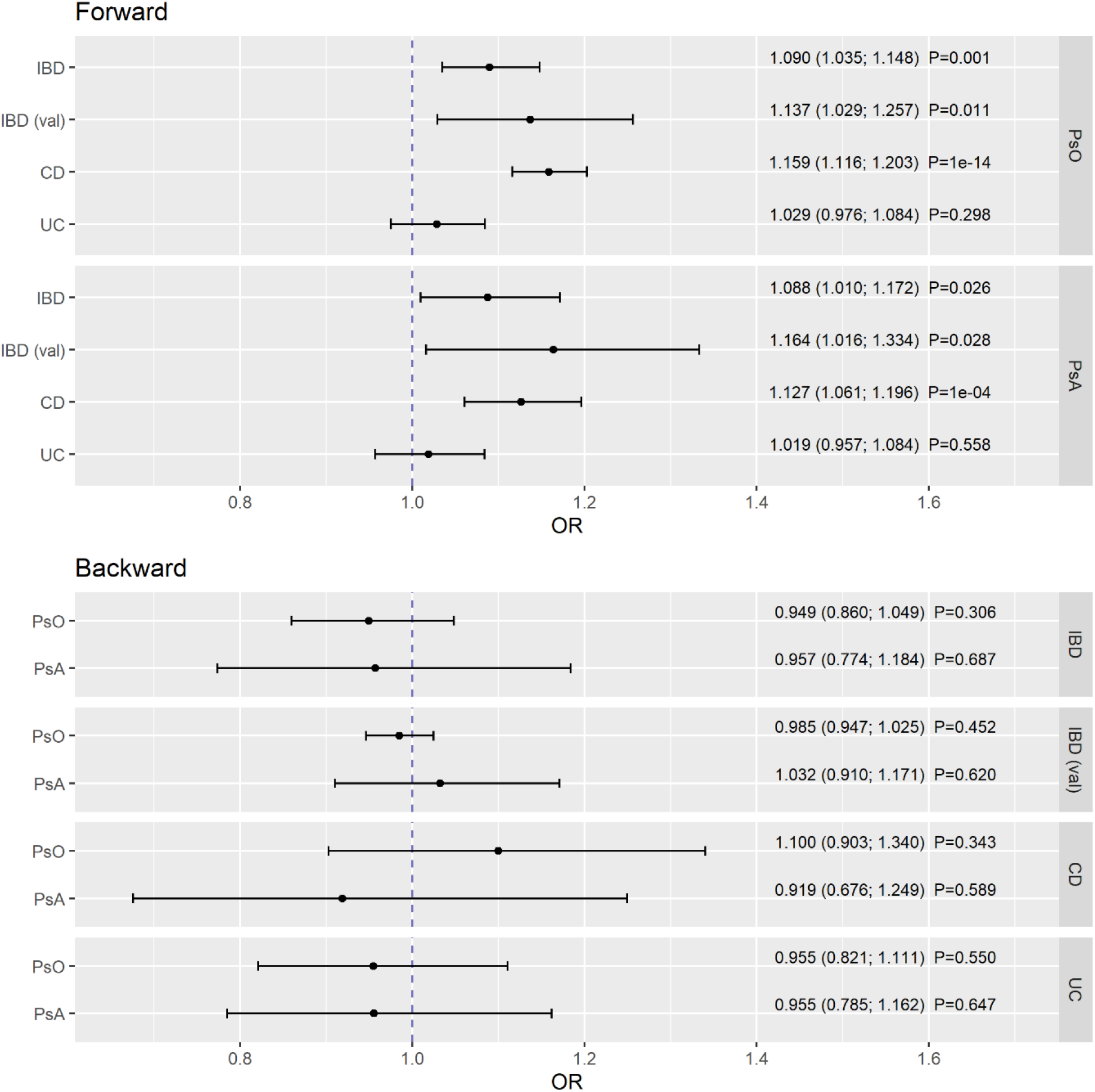
Causal estimates for the impact of inflammatory bowel disease (IBD) and its subentities Crohn’s disease (CD) and ulcerative colitis (UC) on psoriasis (PsO) and psoriatic arthritis (PsA) (forward direction) and vice versa (reverse direction). Estimates are presented as odds ratios (ORs) and 95% confidence intervals from bidirectional Mendelian randomization analyses.

**Figure 2.**
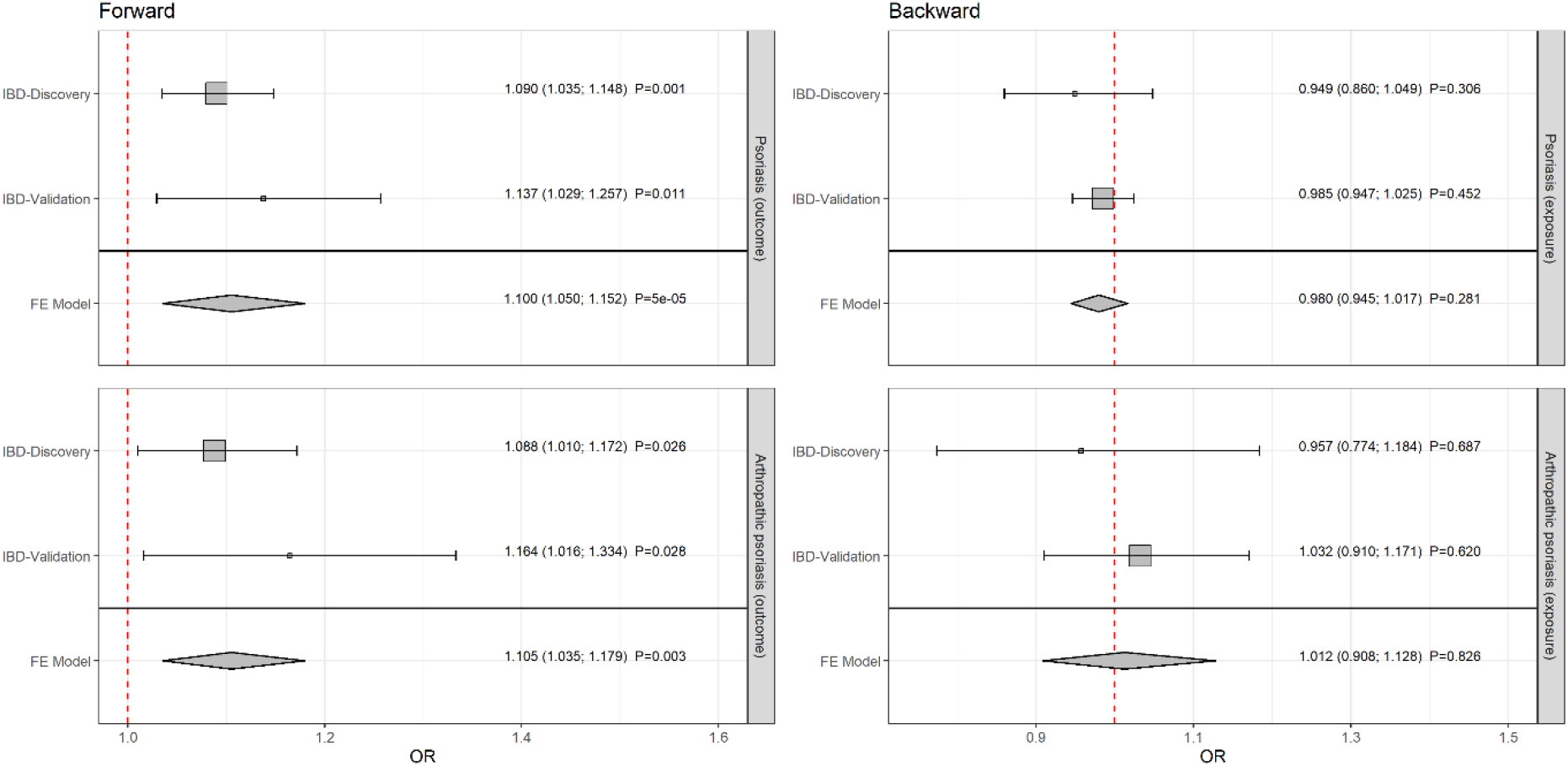
Pooled causal estimates for the impact of inflammatory bowel disease (IBD) on psoriasis and psoriatic arthritis (forward direction) and vice versa (reverse direction). Estimates were calculated applying a meta-analysis fixed effect model on the discovery and validation samples. Results are presented as odds ratios (ORs) and 95% confidence intervals from bidirectional Mendelian randomization analyses.

#### Multivariable MR

The estimates presented for the main results in this section originated from the multivariable robust IVW model with multiplicative random effects. After reciprocal adjustment CD was related to PsO (*OR*=1.16, CI: 1.10; 1.23, *P*= 1 ⋅ 10^−8^) and PsA (*OR*=1.12, CI: 1.04; 1.21, *P*= 2 ⋅ 10^−3^) while UC was not associated with both psoriasis outcomes (PsO: (*OR*=0.98, CI: 0.92; 1.04, *P*=0.414), PsA: (*OR*=0.98, CI: 0.91; 1.07, *P*=0.704)) [Figure 4]. Consequently, the direct effects were even stronger than the total effects from the univariable analyses. Again, no notable genetically predicted associations were observed in the reverse directions [Figure 4].

#### Sensitivity analyses

In the univariable setting, estimates representing the effect of IBD and its subentities on PsO as well as PsA were basically supported by the results of the pleiotropy-robust methods with similar and thus consistent point estimates, indicating reliable results [Supplementary Figures 1 and 2]. However, the MR-Egger point-estimates were somewhat more heterogeneous compared to the other methods. In the reverse directions there was a comparable situation with estimates around the null hypothesis. Apart from that, the MR-RAPS and PRESSO approaches could not be calculated for the effect of CD on PsA regarding numerical issues in consequence of the small number of instruments. No directional pleiotropy could be observed due to the radial MR-Egger intercept test [Supplementary Table 3]. However, some heterogeneity was revealed by the PRESSO global test and the heterogeneity statistics, especially in the models with PsA as exposure.

In sensitivity analyses of the multivariable setting, all pleiotropy-robust approaches supported the main results [Supplementary Figures 3 and 4]. Although there was no evidence for directional pleiotropy, heterogeneity between the particular causal estimates could be observed in the models [Supplementary Tables 4].

Using the results from PhenoScanner search based on genetic instruments for IBD and its subentities [Supplementary Table 5], the network analysis revealed, among others, 3 clusters that were considered as potential confounding factors [Figure 3]. An obesity related cluster consisting of 6 SNPs (red colored), an allergic disease related cluster (16 SNPs; pink, orange, and light blue colored), and an autoimmune disease related cluster (7 SNPs; blue-green colored). Successive removal of the cluster-specific instruments did not substantially change the estimates and thus supported the findings of the univariable as well as multivariable analyses [Supplementary Figures 5 and 6].

The illustration summarizing the results of this study can be found in Figure 5.

**Figure 3.**
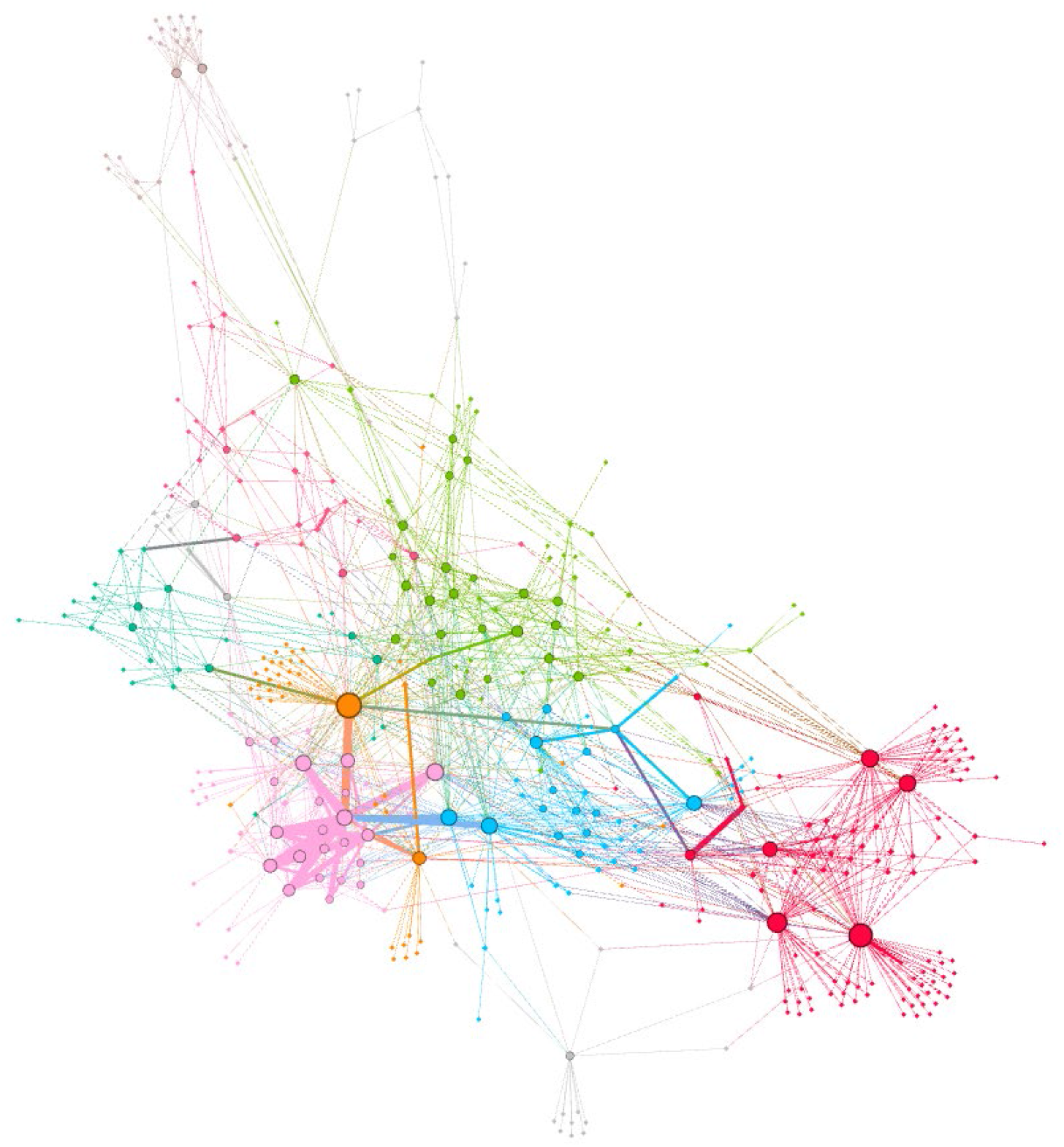
Result from a network analysis showing the associations between genetic instruments and all known phenotypes obtained from a PhenoScanner search.

**Figure 4.**
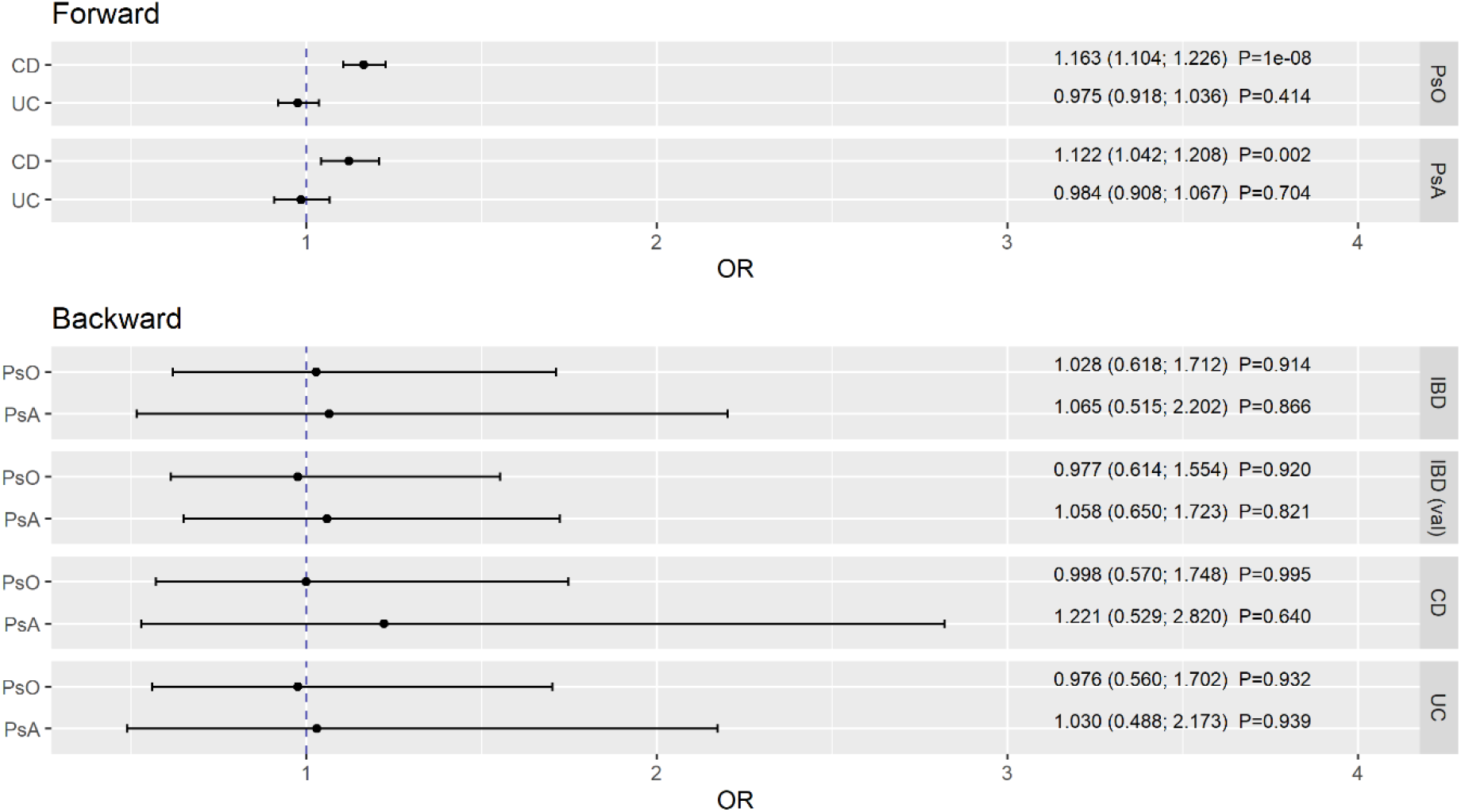
Direct effect estimates obtained from bidirectional multivariable Mendelian randomization analyses. Crohn’s disease (CD) and ulcerative colitis (UC) were mutually adjusted in the forward direction and psoriasis (PsO) and psoriatic arthritis (PsA) in the reverse direction, respectively. Estimates are presented as odds ratios (ORs) and 95% confidence intervals.

**Figure 5.**
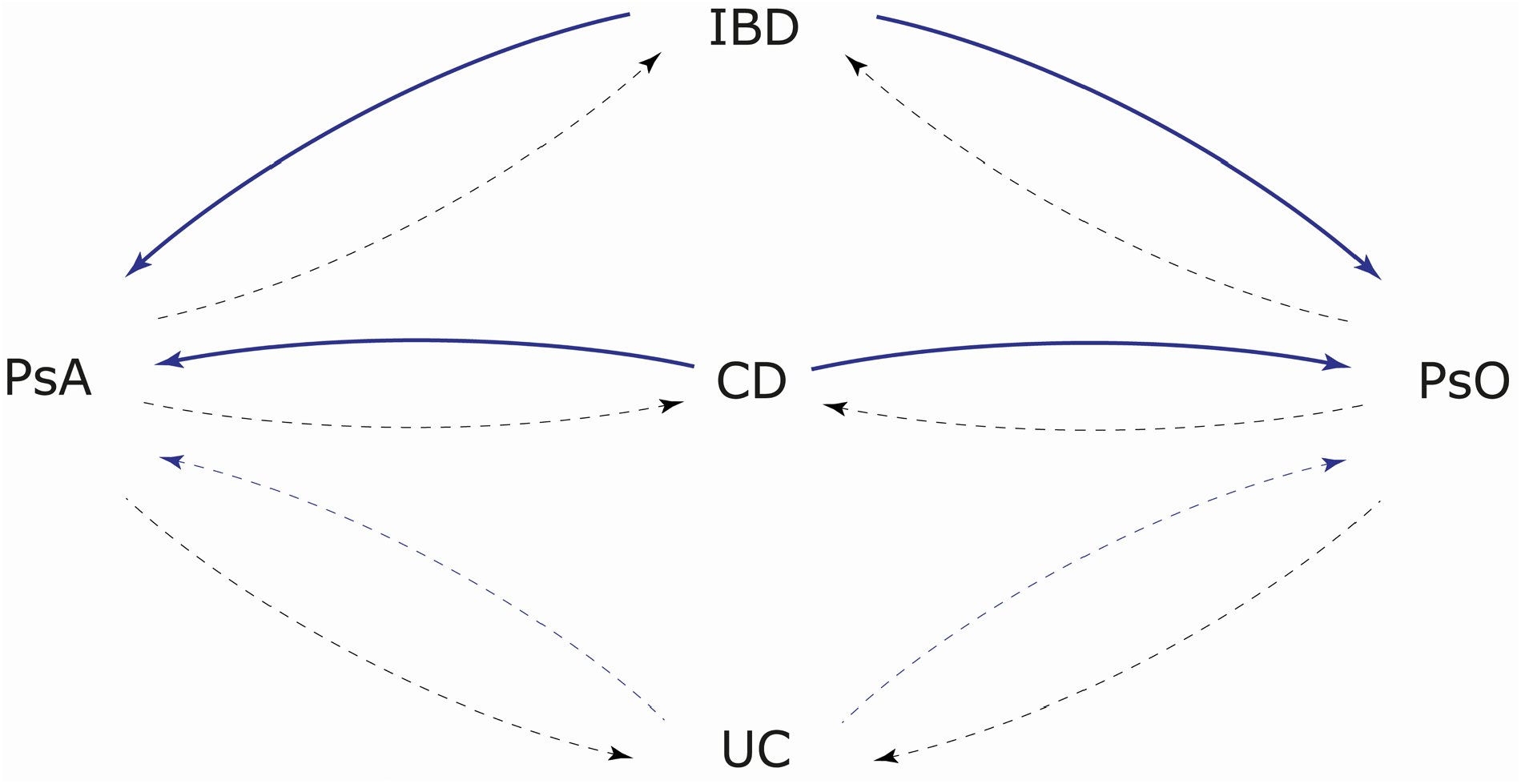
Summary of this Mendelian randomization study. Solid lines depict evidence for a cause-effect chain, while dashed lines indicate no causal associations.

## Discussion

Based on the findings of the present study there is evidence to support a causal link between IBD and PsO and PsA but not vice versa. In particular there was a strong association between CD and both psoriasis outcomes. The results were supported by a series of sensitivity analyses.

Some previous epidemiological cohort studies (18–20) and meta-analyses (21, 22) postulated an association between PsO and IBD. For example, in a nation-wide cohort study from Denmark (n= 5,554,100 individuals aged > 18 years; 75,209 incident cases of PsO, 11,309 incident cases of CD, and 30310 cases of UC) significant relationships between PsO and CD and UC were observed (18). Another nationwide population-based matched cohort study from Korea reported a more than 2-fold increased risk of PsO and PsA in individuals with IBD (20). In a systematic review and meta-analysis including five case-control or cross-sectional studies and four cohort studies with a total of 7,794,087 participants an increased risk and odds of IBD, CD, and UC in patients with PsO was found (21). The meta-analysis based on the four cohort studies revealed a 2.53-fold increased risk of CD and a 1.71 increased risk of UC in patients with PsO in comparison with controls. An increased risk of CD and UC was also observed in patients with PsA (21). Another systematic review and meta-analysis including 93 studies found a significant association between PsO and IBD and vice versa (22). While less than 1% of patients with PsO had CD or UC, the proportion of CD or UC patients with PsO was 3.6% and 2.8%, respectively (22). The prevalence of PsO was highest among children and adolescents with IBD, in particular in subjects with CD. However, heterogeneity between the studies was high (I^2^ ≥ 98%) and a risk of publication bias was likely.

The present study extends previous research by demonstrating a causal effect of IBD, in particular the subtype CD, on PsO, but no causal link in the other direction. The pathophysiologic mechanisms underlying this relationship are not fully understood, but there is little doubt that IBD and PsO likely share a common pathogenesis. Explanations for the association between the two diseases include shared genetic susceptibility loci (16), immune dysfunction (15, 17), gut microbiota dysregulation (34–36), and environmental factors (37). Some GWASs confirmed previously known shared risk loci and identified a number of non-HLA susceptibility loci common to PsO and CD (38). Gut dysbiosis, a state of microbial imbalance, is frequently observed in both IBD and PsO (39). It is well known that diseases of the gastrointestinal tract are often associated with skin manifestations, and the gut microbiome appears to be involved in the pathophysiology of many inflammatory diseases (40, 41). One reason for this might be, that gut microbiota influences epidermal differentiation signaling pathways, thereby affecting skin homeostasis (34, 36). Furthermore, due to intestinal dysbiosis, intestinal permeability may be increased, allowing intestinal bacteria or their metabolites to enter the bloodstream and skin, triggering PsO pathogenesis (34, 35). Also, IBD and PsO share similar immunologic mechanisms. IBD has been shown to lead to an overproduction of Th1 cytokines, with a central role attributed to a class of memory T cells characterized by the presence of cutaneous lymphocyte antigen (CLA) on their surface, which are responsible for settling in the skin and involved in the initiation of PsO (42, 43). In addition, the cytokine TNF-alpha produced by Th17 cells plays an important role in the pathogenesis of IBD and PsO (15).

A strength of this study is that it is the first to examine the causal bidirectional association between IBD including both CD and UC subgroups and PsO as well as PsA using a two-sample MR analysis. In contrast to observational studies, this method of analysis is less prone to confounding, reverse causality, and non-differentially measured exposures with error (44). Using a more conservative, iterative approach, we were able to minimize heterogeneity, confirm consistency of point estimates before and after removal of outliers, and thus strengthen the evidence. A series of sensitivity analyses confirmed the present results. Our findings do not appear to be affected by pleiotropy, as consistent results were obtained in the sensitivity analyses and few outliers were identified using both the iterative IVW and MR-PRESSO methods. The results of this study are based on data from subjects of European descent, and therefore generalizability to other ethnicities is limited.

In conclusions, the present study supports a causal association between IBD and PsO as well as PsA. In particular, the subentity CD seems to be related to the development of both PsO and PsA. Dermatologic consultation is indicated for patients with inflammatory bowel disease presenting with dermatological reactions to initiate a systematic diagnosis, interdisciplinary treatment and management of patients at an early stage. The investigation of pathophysiologic pathways associated with the development of PsO in patients with IBD should be subject of further basic research.

## Supporting information

All supplementary Tables and Figures

## Data Availability

Summary level data for the IBD-discovery (including CD and UC) and IBD-validation samples are freely available from the International Inflammatory Bowel Disease Genetics Consortium (https://www.ibdgenetics.org/downloads.html) and cnsgenomics (https://cnsgenomics.com/content/data), respectively. Instructions for downloading the PsO and PsA data can be found at the FinnGen consortium (https://www.finngen.fi/en/access_results).

https://cnsgenomics.com/content/data

https://www.finngen.fi/en/access_results

## Ethics approval

Not applicable, since the study is based on summary-level data. In all original studies, ethical approval and participant consent to participate had been obtained.

## Author approval

All authors have been personally and actively involved in the substantive work leading to the report. Submission of the manuscript has been approved by all authors.

### Abbreviations

CI: Confidence Interval
GWAS: Genome-Wide Association Study
IBD: Inflammatory Bowel Disease
IVW: Inverse-Variance Weighted
MR: Mendelian Randomization
OR: Odds Ratio
PsA: Psoriatic Arthritis
PsO: Psoriasis
SNP: Single Nucleotide Polymorphism

